# Advancing Family Medicine in a Model Unit: A Living Lab for Health Care Design and Innovation

**DOI:** 10.1101/2024.10.14.24315467

**Authors:** Margaret M. Paul, Marc R. Matthews, Gerry B. Greaney, Deanne W. Wallenstein, Jason D. Greenwood, Tony Spaulding, Jon S. Eckdahl, David R. Rushlow

## Abstract

**Introduction:** The population in need of primary care is rapidly growing and increasingly complex with respect to chronic disease burden. We must develop alternative and more efficient approaches to managing patients if we are to increase access to care without sacrificing continuity; however, there is little guidance for innovation strategies at the practice level.

**Methods:** The Mayo Clinic Department of Family Medicine engaged in a 2-year multistage planning process to develop plans for a Model Unit (MU) to identify opportunities for innovation to improve daily practice. The purpose of the MU is to operate as a “living lab” capable of driving continuous advancements within the context of a real-world health system with the goal of delivering high-quality care to a greater number of patients.

**Results:** Key lessons from the planning stage led to the development of a contextualized, incremental, and continuous approach to design and innovation. In its first phase, the MU includes 3 interventions that are novel to the unit itself, including nurse-led hypertension management, incorporating telehealth visits into routine clinician schedules, and ambient documentation to replace clinician-generated visit notes. We present a description of the overall MU approach including early-stage implementation findings and our evaluation strategy.

**Conclusions:** The MU structure is a generalizable model for identifying opportunities and operationalizing practice improvement activities in a strategic and pragmatic way that incorporates real-time feedback from clinicians and staff with the expectation of continuous and phased evolution.

## Introduction

The population in need of preventive care and chronic disease management is rapidly growing. The number of people aged 50 years and older will increase from 137 million in 2020 to 221 million by 2050, with chronic disease burden increasing at twice the rate, from 72 million to 142 million over the same period of time.^1^ Concurrently, there is a deficit of primary care physicians that is estimated to grow to a national shortage of 139,160 physicians by 2030.^2^ The primary care workforce is already feeling the effects of this burden; physicians and staff are stressed, with increasing rates of dissatisfaction and burnout widely reported throughout the country.^3,4^ These problems are not new, but they continue to worsen and fuel the problem of a dwindling supply of primary care physicians.

Panel size is often at the center of discussions on how to improve physician satisfaction, and smaller panel sizes have indeed been associated with higher quality of care.^5^ Recent research suggests that primary care panel sizes may actually be shrinking, but at the expense of access to care at the population level.^6,7^ We must develop alternative and more efficient approaches to managing patients if we are to increase access to care without sacrificing continuity. Advances in health care related to artificial intelligence (AI), other digital tools, and expanded access and use of telehealth have unlocked potential to change the way we practice medicine; however, more research is needed to assess strategies to implement these structures and processes in real-world practice settings, and their impacts on access, continuity, quality, and outcomes.

The Mayo Clinic Department of Family Medicine has developed a Model Unit (MU) to evaluate new approaches to care in the context of its large integrated practice in the upper Midwest. The purpose of the MU is to operate as a “living lab” capable of driving continuous advancements within the context of a real-world health system with the goal of delivering high-quality care to a greater number of patients while working within the constraints of limited staffing. The fundamental assumption of the Unit and the interventions tested therein is the hard truth that providing access and care continuity to our community means that panel sizes will inevitably grow due to the immense challenge of recruiting enough clinicians (physicians and advanced practice providers) and allied health staff to meet the rising demand for care, both now and in the future. Importantly, the MU is not meant to have a final goal, to implement a specific care model, or to only experiment with novel technologies. Rather, it proposes a new way of designing and running practice improvement activities in a practical way that integrates real-time feedback from clinicians and staff with the anticipation of nimble, continuous, and gradual change.

The MU approach to localized design and innovation was developed in response to existing models of care, such as the chronic care model ^8,9^ and the patient-centered medical home ^10^, which require broad and sweeping transformation to daily practice operations, and are often implemented at a health system level. In contrast, the MU approach is incremental, and “bottom-up”, with innovation driven by thoughtful design that is guided by the practice in which the changes are implemented and is focused on the role of discrete team structures and processes in influencing positive changes in health care quality and clinical outcomes.^11^ It is distinct from continuous quality improvement programs,^12^ which are focused on adherence to existing standards and best practices, and from practice transformation efforts in general, which aim to achieve specific objectives and therefore have predetermined end points in which a new stable state is achieved. It is also distinct from operationalizing Lean principles^13^ because it is focused on facilitating ongoing practice-level, and practice-driven, changes as opposed to adherence to an overall guiding structure for organizational management and transformation.

While facets of each of these approaches to practice improvement may inform various interventions evaluated in the MU over time, it is a demonstration of a unique approach to driving contextualized innovation and transformation within a large health system.

Our Unit is based on four core principles which support the focal point of our model: practice-driven design to facilitate transformative innovation. (Figure 1) In the context of the MU, health care design is: 1) contextualized and originates from within the practice area of focus; 2) collaborative in that its function is dependent on insights and action from a multidisciplinary and dynamic team; 3) a continuous cycle of evaluating our interventions, responding to feedback loops with clinicians and staff, and allowing innovations to adapt and evolve to meet the needs of the practice; and 4) dependent on iterative, strategic prototyping to examine new ideas in the context of a real-world clinic.

**Figure 1.**
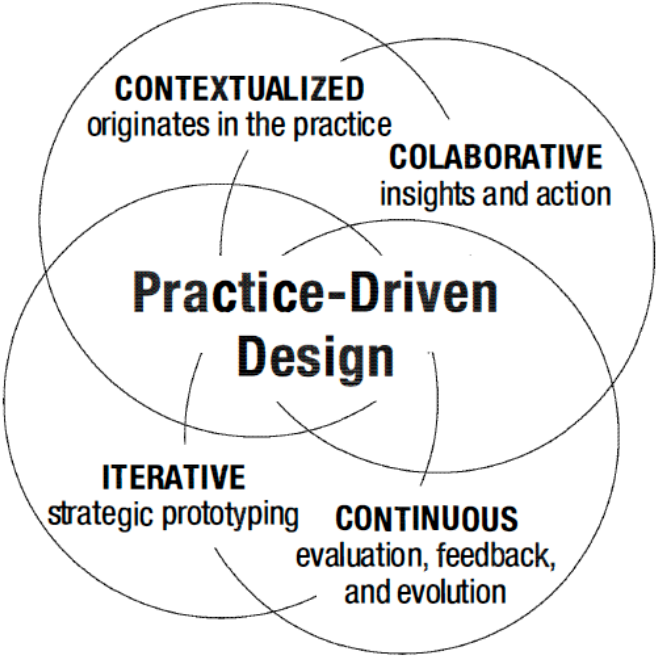
Core Innovation Principles of the Model Unit

These principles ensure that the individual interventions implemented through the MU approach are highly dependent on the context of our specific practice; however, the structures and processes required to plan, implement, and evaluate the activities of the MU are generalizable to other large health systems. In this paper, we introduce the overall structure of the MU, share our plans for the first three phases and related research and evaluation, and our vision for sustainability. In addition to assessing individual interventions, we describe our structures to facilitate ongoing assessment of the MU overall to monitor its impact on the practice and adapt as necessary to support acceptability to a diverse group of stakeholders and sustainability.

## Methods

Planning for the MU began in 2021 with the formation of a multidisciplinary stakeholder committee based at the Mayo Clinic in Rochester. The group included clinical leadership representation from the Department of Family Medicine as well as clinical, informatics, nursing and administrative leadership from the clinical site itself. The purpose of this committee was to explore opportunities for innovation in the Mayo Midwest practice, a network of 56 primary care practices located throughout parts of southeastern Minnesota, western Wisconsin, and northwest Iowa. An initial phase of this work focused on broad scale, system-wide implementation of bundled interventions to improve family medicine practice throughout Mayo Midwest locations utilizing our pre-established department governance structure. Rather than test interventions on a system level, which has proven difficult to implement and research, the stakeholders decided to focus on piloting new interventions within a single team in a Rochester-based family medicine clinic. The selected care team is diverse and has a long history of innovation and dedicated support and vision from physician leaders within the practice, therefore the stakeholder committee felt that they were a superb choice for the MU, capable of readily transforming daily practice to test innovative approaches to care.

In spring 2023, after identifying the team that would become the MU, the stakeholder group was transitioned by two new committees focused on operationalizing their vision: the Design Committee and the Research Committee. The Design Committee is the governing body of the MU and is responsible for engaging the care team in defining the overall structures of the MU, selecting interventions, monitoring implementation progress, and identifying primary and secondary outcomes for each intervention. This team, much like the prior stakeholder committee, is multidisciplinary and includes clinicians and staff practicing in the MU, clinical informatics specialists, family medicine department leadership, experts in strategy and service design, and health services researchers. The MU Research Committee is a subgroup of the Design Committee responsible for planning the details of the evaluation and dissemination efforts related to MU planning, implementation, and evaluation.

In fall of 2023, the Design Committee determined the initial process for identifying, implementing, and testing structures and processes in the MU. We decided to use a phased approach which includes discrete cycles to implement and evaluate short-term outcomes, bundling 2-3 interventions per cycle. Over the course of about 6 months, the Design Committee continued to refine the structure and overall goals of the MU and operationalized our innovation principles into a framework that also represents our core structures (Figure 2). The Unit is built upon the rich history and culture of innovation of Family Medicine at Mayo Clinic and the underlying assumptions that our success depends on our ability to collect continuous feedback and communicate with stakeholders about our work. Prototypes are at the heart of our design process and are developed with input from the Design and Research Committees, the MU care team broadly speaking, from physicians to desk staff, and patients as appropriate. Together, our three committees are responsible for the overall vision and execution of the Unit, setting priorities that are aligned with those of the Mayo Clinic enterprise, and managing the implementation, evaluation, and dissemination of our findings.

**Figure 2.**
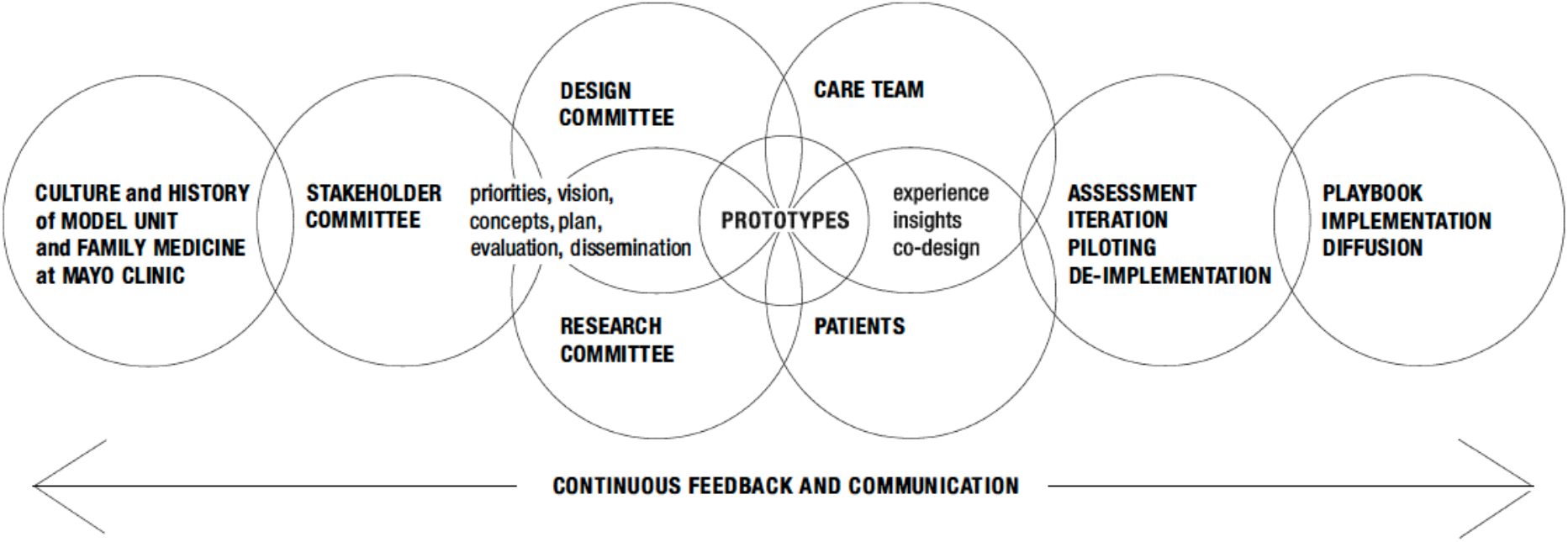
The Model Unit Framework

To date, we have established plans for three phases of the Unit, and our plan is to extend this work indefinitely through this phased approach. The first phase, currently underway, is focused on internal operations to enhance our operational efficiency within the practice. The interventions for phase 1 were selected to address three critical areas of focus defined by the Design Committee, including improving chronic disease management, increasing flexibility in scheduling for patients, clinicians, and staff, and reducing clerical and cognitive burden among clinicians and staff. Our second phase will focus on pre-visit communication between the care team and our patients with the goal of improving continuity through patient engagement. In phase 3, we will develop interventions to create and implement care and visit pathways that expand visit request end points beyond a one-on-one, in-person visit with the provider to better match patients to the appropriate provider and visit type (i.e., in person or virtual) based on their needs.

## Results

### Model Unit Structure

The MU is located at the Mayo Clinic in Rochester and is co-located on the same campus as multiple specialty clinics with other services, such as lab, imaging, and pharmacy. The MU team is composed of 6 physicians, 5 advanced practice providers (APPs), 8 nurses, and 1 scheduler who collectively care for 10,506 patients annually. The current average risk adjusted panel size is 1,979 (range 1,124 to 2,940) patients per physician and 825 patients (range 644-1,044) per APP.

### Phase 1 Interventions

In its first phase, we implemented 3 interventions designed to address the areas of focus defined by the Design Committee to cut across all phases of the MU: to improve chronic disease management, reduce clerical burden, and increase flexible scheduling (Table 1). Each intervention has at least one primary outcome associated with it which we felt could reasonably show effects within 9 months of implementation.

**Table 1.** Model Unit Phase 1 Interventions and Measures of Success.

| Interventions | Key Outcomes | Goals | Timing |
| --- | --- | --- | --- |
| **Nurse-led hypertension management combined with new PSA role to create slack for panel management.** | proportion of patients with nurse-led hypertension management | 20% of eligible patients under nurse management | 6 months |
|  | patient blood pressure | improvement in blood pressure over time for newly empaneled patients AND no difference in trajectory of BPs for patients under clinician vs nurse-led management | 1 year |
|  | increased complexity of physician patient panels | increased complexity of physician panels as measured by increase in average panel risk score | 9 months |
|  | decrease in average # of patients roomed by nursing staff in each week | 50% decrease in RN rooming | 6 months |
| **Ambient Documentation** | adoption (proportion of eligible clinicians using it) | 100% of clinicians with access are using the tool | 3 months post-implementation |
|  | Time dedicated to documentation (minutes/day) and cognitive burden from generating visit notes | Reductions in documentation time and cognitive burden associated with note taking | 3 months post-implementation |
| **Regular use of telehealth visits** | appointments available | 50 slots available (5/week/physician) | 6 months |
|  | appointments filled/available | 80% completed appointments of those appointments made available | 6 months |
|  | ratio of telehealth to in-person visits | gradual increase to every provider having 1/2 day of telehealth appointments each week | 6 months |

Nurse-led hypertension management will be implemented to improve chronic disease management and facilitate staff and providers to work at the top of their skillsets. The nurse and physician partners on the Design Committee highlighted the challenge in task shifting from clinicians to nurses, namely that the nurses could not reasonably pick up hypertension management without being able to shift some of their clerical tasks to other staff members, and so the MU will add three unlicensed Procedural Support Assistants (PSA), to room patients and support clinic operations activities to off-load this task from nursing staff. The primary objective for this intervention is to increase the number of patients under nurse-led management for hypertension without sacrificing care quality or outcomes so that physician time can be redirected at patients with undiagnosed, more serious or complex conditions without a treatment plan in place. In addition to these key measures, we will also monitor trends in blood pressure control among nurse vs physician led patient panels to ensure that our standards of care are maintained, as well as the amount of rooming done by nurses vs PSAs to ensure that the planned task shifting is occurring in daily practice.

To decrease the substantial clerical burden of documenting visit notes, the MU is testing a new technology, generally described as ambient documentation, which utilizes a generative AI vended platform to auto-create the clinician note during the visit. While the workflow designed to integrate ambient documentation into clinical practice requires physician review, this task is not nearly as burdensome as creating the note independently. Early reports from clinicians have been extremely positive with a few seeing 80% reduction on documentation time and 90% improvement in same day encounter closure rate. We will continue to monitor use of this tool and qualitatively explore barriers to use. The primary outcome for this intervention is reduction of clinician clerical and cognitive burden with the possibility of improving access.

Finally, to increase flexibility in physician workplace, reduce stress on allied health staffing, and improve facility utilization, the MU will incorporate telehealth visits into routine scheduling with the goal of each physician having one half-day a week of video visits by the end of the 6-month cycle. Physicians can choose to perform these visits from home on a secured/encrypted laptop device. Early findings suggest that this may increase efficiency in the clinic (i.e., less prep time for staff per patient), reduced utilization of exam rooms and on current facilities, and increase clinician satisfaction. At the end of the first phase of testing, we hope to see an increase in the availability and fill rates of telehealth appointment slots.

## Discussion

The primary objective of the MU is to identify effective interventions that allow us to redefine the way we manage patient panels to allow us to provide better health care to a greater number of patients within the very real constraints of day-to-day clinical practice. The core underlying assumption of the MU is that there are ample promising and feasible opportunities for practice improvement available to optimize practice efficiency and improve patient care quality and continuity. The purpose of the structures developed to support the MU are to identify such opportunities and select them for implementation and to evaluate the implementation and outcomes of the selected interventions. Any effective strategies can then be implemented on a wider scale whereas ineffective interventions will be not be adopted by the practice.

Lessons from the MU planning stage led to the development of a pragmatic, phased and continuous approach to innovation. The inclusion of MU team members, including clinicians and nurses, has enhanced our design processes by increasing the feasibility of our plans. In particular, these committee members have highlighted the need for creating the bandwidth required in each phase to make room in existing workflows for new interventions. Indeed, this has been one of the most challenging tasks for the Design Committee to date, as it gets at the crux of the challenge we are trying to address through innovation: how to provide better care with the same amount of staff resources. Our diverse stakeholder group has also highlighted the need for development of innovative measures of success on the business side of the practice as we implement innovations that shift away from traditional goals such as outpatient visits.

Leadership from our Design Committee have begun regular meetings with leaders in finance and administration to determine how best to define success over the next year of productivity in the MU and beyond.

The rapid, phased nature of the MU structure has the potential to contribute to burnout among MU staff and clinicians. Therefore, in terms of evaluation, we rely to the extent possible on existing data which can be extracted from the electronic health record (Epic) and other administrative datasets that are automatically generated through routine clinical care. While collecting data on proximal measures of success, such as clinician and staff satisfaction with each specific intervention, would be helpful, it is not practical to collect the primary data needed to ascertain such measures. However, we plan to occasionally conduct more in-depth evaluations of specific interventions that are of particular interest to the Design Committee and other stakeholders. In these instances, we will employ mixed methods including surveys, interviews and EHR data analysis with matched comparison groups to better elucidate the impact of that specific intervention of interest.

The immediate next steps for the MU teams are to fully implement phase 1 interventions, evaluate their impact, and determine which will be retained in routine care. Phase 2 planning is already underway, and the next round of interventions will be implemented in late 2024. Within the next year, we hope to expand panel sizes as informed by the quantitative and qualitative data from the initial interventions to better satisfy demand and achieve our goal of providing better care to more patients. Therefore, as with phase 1, the interventions for phases 2 and 3 will be selected in part based on their ability to create slack in clinician and staff schedules. One of the most significant concerns raised by the care team members in the clerical burden generated by the EHR that has been echoed many times by others,^14-17^ and so it is likely that at least one of the phase 2 interventions will be designed to address this critical issue.

## Conclusion

The MU is the Mayo Clinic approach to practice-level, pragmatic, and rapid innovation in family medicine. The MU does not represent a specific, preexisting model of care, nor is it evolving to align with a predetermined vision of ideal practice. Instead, it is a new approach to structuring practice improvement activities and facilitating bottom-up innovation with the expectation of continuous, phased, and contextualized evolution.

## Data Availability

All data produced in the present work are contained in the manuscript.

## Acknowledgements

The authors thank past and current members of the Model Unit Stakeholder, Design, and Research Committees for their contributions to this work.

